# Markerless Video Analysis of Spontaneous Bodily Movements in 4-Month-Old Infants Predicts Autism-like Behavior in 18-Month-Olds

**DOI:** 10.1101/2021.10.11.21264725

**Authors:** Hirokazu Doi, Naoya Iijima, Akira Furui, Zu Soh, Kazuyuki Shinohara, Mayuko Iriguchi, Koji Shimatani, Toshio Tsuji

## Abstract

Early intervention is now considered the core treatment strategy for autism spectrum disorders (ASD). Thus, it is of significant clinical importance to establish a screening tool for the early detection of ASD in infants. To achieve this goal, in a longitudinal design, we analysed spontaneous bodily movements of 4-month-old infants and assessed their ASD-like behaviours at 18 months of age. Infants at high risk for ASD at 18 months of age exhibited less rhythmic and weaker bodily movement patterns at 4 months of age than low-risk infants. When the observed bodily movement patterns were submitted to a machine learning-based analysis, linear and non-linear classifiers successfully predicted ASD-like behaviour at 18 months of age based on the bodily movement patterns at 4 months of age, at the level acceptable for practical use. This suggests the utility of the proposed method for the early screening of infants at risk for ASD.

## 1. Introduction

Autism spectrum disorder (ASD) is a developmental disorder, typically characterised by a collection of symptoms, including repetitive behaviours, restricted interests, and poor social communication skills (American Psychiatric Association, 2013). ASD is generally considered to have genetic underpinnings (Anderson, 2015). Recently, the prevalence of ASD has been increasing. This is partly owing to increased public awareness and changes in the ASD diagnostic criteria. However, a significant increase in the prevalence of ASD remains unexplained; hence, there is a pressing need for establishing reliable treatment strategies for ASD.

At this point, there is no definitive cure for ASD, but an increasing number of studies indicate the efficacy of early intervention for children with ASD (Dawson et al., 2010; Green et al., 2015), which is manifested in improved prognosis and social adaptation. Thus, early detection of children at risk for ASD and early intervention are now considered to be a core treatment strategy for ASD (Dawson, 2008).

It is generally accepted that ASD can be diagnosed at around 3 years of age. However, a number of studies have indicated that early signs of ASD can be detected during infancy and toddlerhood (Adrien et al., 1993; Baranek, 1999; Osterling, Dawson & Munson, 2002; Klin et al., 2009). For example, a retrospective study by Osetrling et al. (2002) analysed birth-day videos of ASD and typically developed (TD) children, and found that children later diagnosed with ASD showed lower social functioning compared with TD children. Likewise, a prospective study by Elsabbagh et al. (2012) revealed that children with ASD show atypical electrophysiological activation to socially significant facial information (e.g. direct gaze), compared with their siblings without ASD and with TD children as young as 10 months of age.

Previous findings pertaining to the early signs of ASD raise the possibility that children at risk for ASD can be screened during toddlerhood or, in some cases, during infancy. Early detection could lead to early intervention, potentially improving the prognosis for children with ASD and/or for those with sub-clinical-level symptoms. However, presently, there are no established tools for early detection of the ASD risk, and screening of children at risk for ASD requires time-consuming assessments and/or observations by teams of multidisciplinary professionals consisting of experienced clinicians and psychologists (Zwaigenbaum et al., 2009).

Atypical motoric function is one of the most frequently reported signs of ASD during early development (Teitelbaum et al., 1998; Ozonoff et al., 2008). Many studies have reported various types of atypical bodily movement patterns, from infancy and toddlerhood, such as unbalanced posture and movement (Esopsito et al., 2009, 2011; Dawson et al., 2018), hypotonia (Adrien et al., 1993), spasticity in limb extremities (Dawson et al., 2000) and impairment in manual movement control (Sacrey et al., 2018). These behavioural observations are in line with neuroimaging-based findings and histological studies on the brains of humans with ASD, in which atypical patterns have been reported regarding the morphology and function of motor-related neural regions, such as the basal ganglia and cerebellum (Rinehart et al., 2006; Qiu et al., 2010).

In studies on motor function during infancy, intensive attention has been paid to spontaneous movements in the supine position, which can be evaluated starting from the neonatal period. Neonates and infants exhibit a repertoire of bodily movement patterns in the supine position, which has been termed the general movement (GM). Prechtel et al. investigated the developmental trajectory of GM in TD children and proposed a framework for qualitative assessment of GM that can be used for predicting the risk of deficiency in higher-order brain functions, such as cerebral palsy (Einspieler et al., 2016; Burger & Louw, 2009). Although the number is relatively small, several studies have also indicated the utility of GM for assessing the risk of psychiatric conditions (Einspieler et al., 2014; Hadders-Algra, Bouwstra & Groen, 2009).

Considering the success of the GM assessment for predicting the risk of neurological and psychiatric conditions, together with the prevalence of atypical motoric development patterns in children with ASD, it seems feasible to establish an early screening method for objectively evaluating the risk of ASD based on the analysis of bodily motion patterns during early development. Several researchers have developed systems for automatic assessment of GM using image analysis techniques (Gao et al., 2019; Marcroft et al., 2015; Støen et al., 2009). However, these studies did not address the possibility of the ASD risk evaluation.

The primary goal of the present study was to examine whether bodily movement patterns during early infancy are predictive of the ASD risk, which becomes evident later in the development in a prospective design. To achieve this goal, we video-recorded spontaneous bodily movements in 4-month-old infants and quantified their features using a novel markerless system for infant movement evaluation (Osawa et al., 2009; Tsuji et al., 2020; Kinoshita et al., 2020; Kawashima et al., 2020; Tacchino et al., 2021). Classifiers were trained for predicting the ASD risk in 18-month-old infants, based on their bodily movement patterns at 4 months of age. The design of the current study is schematically shown in Figure 1. In contrast to some of the other proposed methods of automated infant movement analysis, our system does not require special equipment or attachment of sensors to the body surface; thus, it can be easily implemented in clinical settings.

**Figure. 1.**
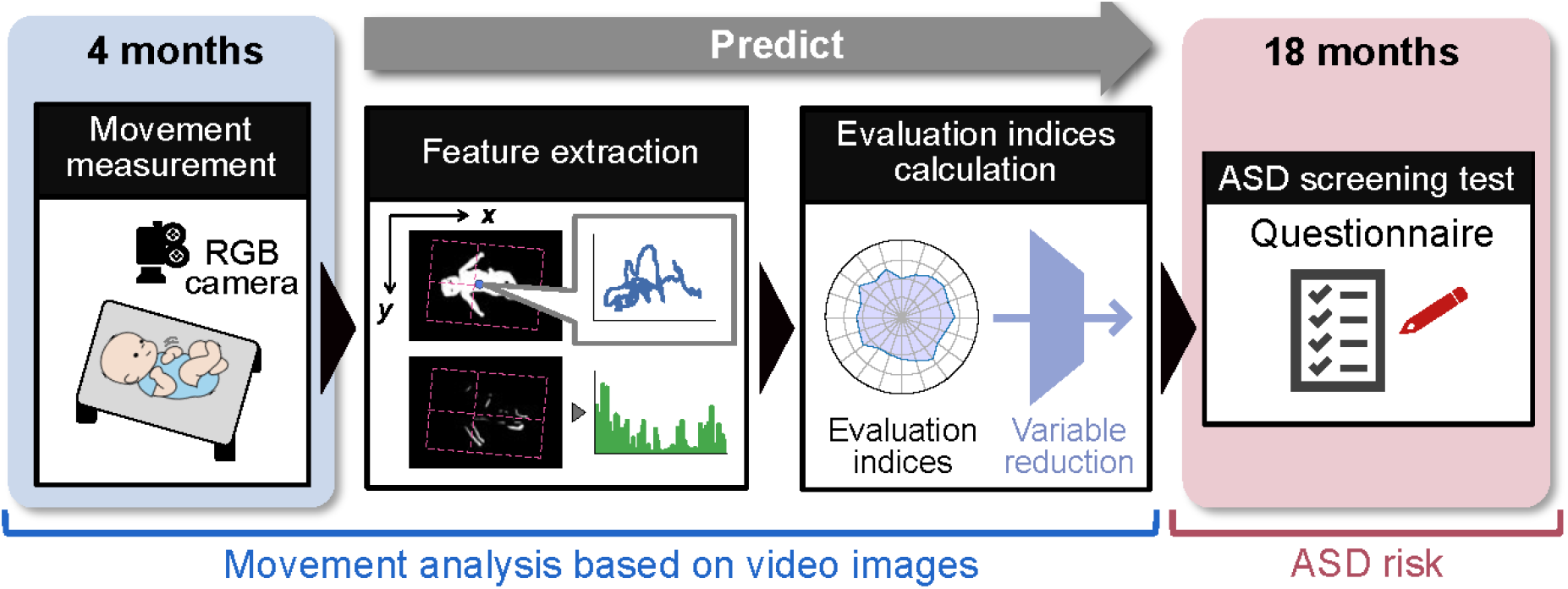
Schematic representation of infant video recording setting and the flow of information processing.

The risk of ASD in 18-month-old infants was evaluated using the Modified Checklist for Autism in Toddlerhood (MCHAT) (Inada et al., 2011; Kamio et al., 2015). The MCHAT is a questionnaire widely used for early screening of the ASD risk, and is reported to be applicable to toddlers as young as 18 months of age. Although the specificity of the MCHAT method is not very high, it captures early signs of atypical development broadly in many domains, such as social communication and sensory processing.

## 2. Results

Of the 62 mother-infant pairs who participated in the video-recording sessions at 4 months of age, 58 mothers returned the MCHAT questionnaire at 18 months of age. Among these 58 mother-infant pairs, bodily movement analysis was performed for 41 infants. Video recordings from 17 infants were discarded owing to their short length (≤ 3 min; *n* = 13) and low frequency (*I*_1_ ≤ 10%; *n* = 4) of bodily movement in them. There were no significant differences between these 41 mother-infant pairs and 21 pairs whose data were discarded in terms of the days after birth (*W* = 1.439, *p* = 0.156), the weight at birth (*W* = 1.227, *p* = 0.227), the gestational age at birth (*W* = 1.032, *p* = 0.308), and mother’s age at birth (*W* = -1.6, *p* = 0.119). The distributions of high-and low-risk children did not differ between 41 mother-infant pairs and 17 mother-infant pairs whose MCHAT data were available but were discarded from the final analysis (*p* = 0.715); this conclusion was reached based on Fisher’s exact test. Thus, although the data attrition rate was relatively high, there were no signs of selection bias.

Among the 41 infants, seven were evaluated to be at high risk for ASD (the high-risk group). The age-in-days on the day of video recording, weights at birth, the gestational ages of the infants, and the mothers’ ages, for the high-and low-risk groups, are summarized in Table 1. The Brunner— Munzel test did not reveal significant group differences either in age (*W* = 0.067, *p* = 0.95), weight at birth (*W* = -1.400, *p* = 0.18), gestational age (*W* = 0.214, *p* = 0.835), or mother’s age at birth (*W* = 1.172, *p* = 0.265).

**Table 1.** Background information of infants and mothers. In the parenthesis are the standard deviations.

| Group | High Risk | Low Risk | $W$ | $p$-value |
| --- | --- | --- | --- | --- |
| Days after birth at recording (days) | 128.6 (10.4) | 125.9 (5.7) | 0.0669 | 0.949 |
| Weight at birth (g) | 2964.0 (221.7) | 3121.2 (383.1) | -1.400 | 0.179 |
| Gestational age (days) | 278.9 (7.8) | 278.1 (7.9) | 0.214 | 0.835 |
| Mother's age at birth (yrs) | 31.3 (3.6) | 33.4 (4.9) | -1.172 | 0.265 |

*2-1. Feature Selection*

### 2-1. Feature Selection

We identified and quantified 26 bodily movement features; these are summarized in Table 2. Appendix contains more detailed definitions of these bodily movement features. The boxplots of each bodily movement feature extracted from video recordings are shown in Figure 2 for the low- and high-risk groups.

**Table 2.** Summary of bodily movement features.

| Feature | Definition |
| --- | --- |
| $^{(A_k)}I_1$ | Movement frequency in body region $k$ ($A_k$; $k = 1 \sim 6$) |
| $^{(A_k)}I_2$ | Movement strength in body region $k$ ($A_k$; $k = 1 \sim 6$) |
| $^{(A_k)}I_3$ | Movement count in body region $k$ ($A_k$; $k = 1 \sim 6$) |
| $^{(A_5, A_6)}I_4$ | Ratio between movement frequency in upper ($A_5$) and lower body ($A_6$) regions |
| $^{(A_5, A_6)}I_5$ | Ratio between movement strength in upper ($A_5$) and lower body ($A_6$) regions |
| $^{(A_5, A_6)}I_6$ | Movement coordination between upper ($A_5$) and lower body ($A_6$) regions |
| $(A_{5,6})I_7$ | Central frequency of time-series of movement in upper ($A_5$) and lower body ($A_6$) regions |
| $(A_{5,6})I_8$ | Second moment around central frequency of time-series of movement in upper ($A_5$) and lower body ($A_6$) regions |
| $I_{9,x}, I_{9,y}$ | Central frequency of time-series of body center velocity along x- (top-bottom) and y- (left-right) axes |
| $I_{10,x}, I_{10,y}$ | Second moment around central frequency of time-series of body center velocity along x- (top-bottom) and y- (left-right) axes |
| $I_{11,x}, I_{11,y}$ | Central frequency of time-series of body center fluctuation along x- (top-bottom) and y- (left-right) axes |
| $I_{12,x}, I_{12,y}$ | Second moment around central frequency of time-series of body center fluctuation along x- (top-bottom) and y- (left-right) axes |
| $I_{13,x}, I_{13,y}$ | Average of absolute values of body center velocity along x- and y-axes |
| $I_{14,x}, I_{14,y}$ | Standard deviation of body center fluctuation along x- and y-axes |
| $I_{15}$ | Averaged area of body center excursion |

**Figure. 2.**
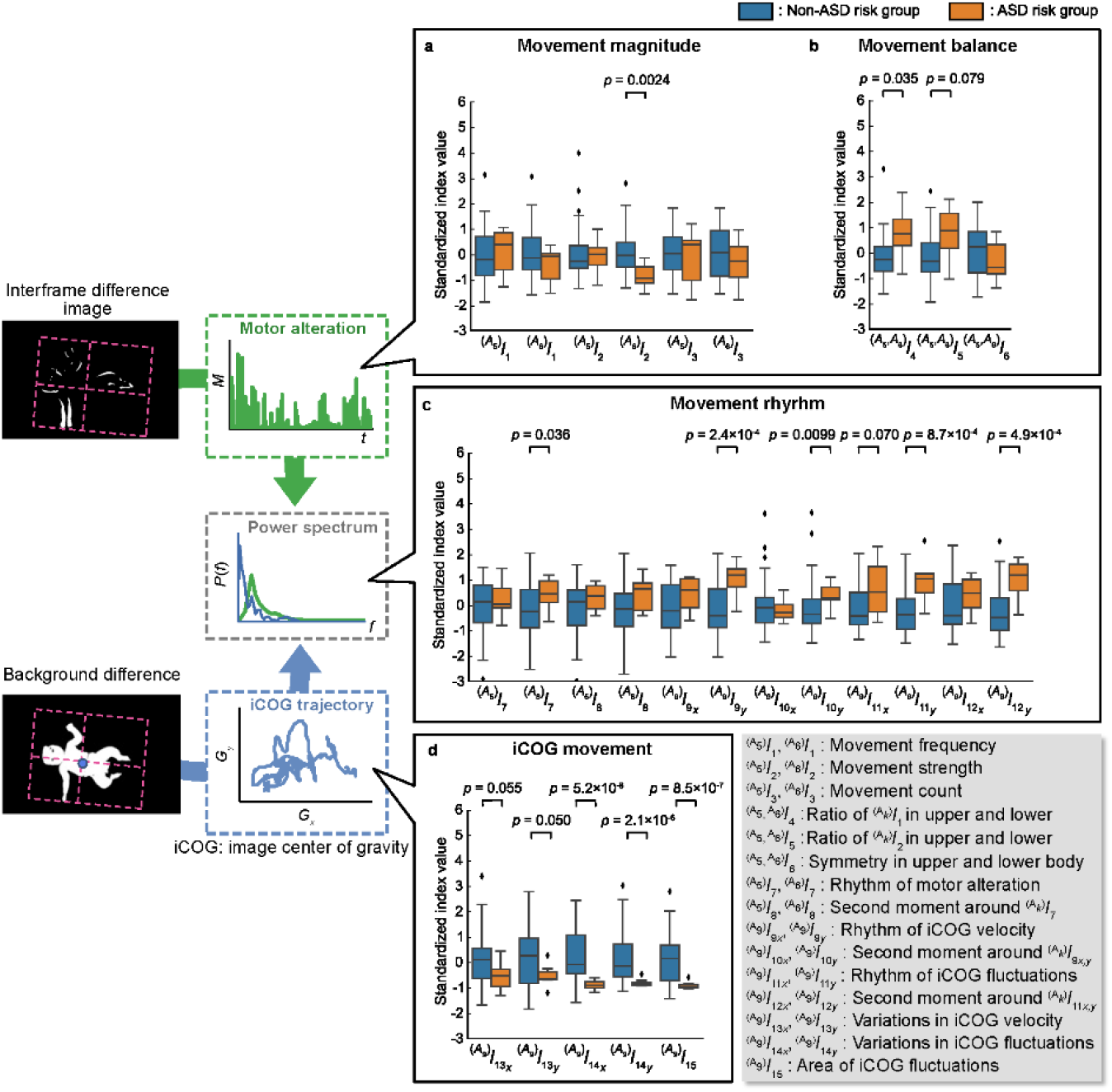
Boxplots of features. a) movement magnitude, b) movement balance, c) movement rhythm and d) movement of the center of gravity (iCOG movement) in each group.

As can be seen from Figure 2, infants in the high-risk group exhibited significantly lower movement strength in their lower limbs (^(A6)^ *I*_2_ ; *W* = -4.10; *p* = 0.0023) and lower balance of movement between their upper and lower limbs (^(A5,6)^ *I*_4_; *W* = 2.56, *p* = 0.035). In the domain of movement rhythmicity, infants in the high-risk group exhibited higher central frequencies and larger second moments around the central frequency in the time series of the body centre velocity, and a fluctuation along the left-right axis (*I*_9, y_, *W* = 4.94, *p* < 0.001; *I*_10, y_ *W* = 2.89, *p* = 0.0099; *I*_11, y_, *W* = 4.17, *p* < 0.001; *I*_12, y_, *W* = 4.76, *p* < 0.001). Examples of the temporal course of body center fluctuation and the distribution of frequency power are shown for low- and high-risk infants in Figure 3.

**Figure 3.**
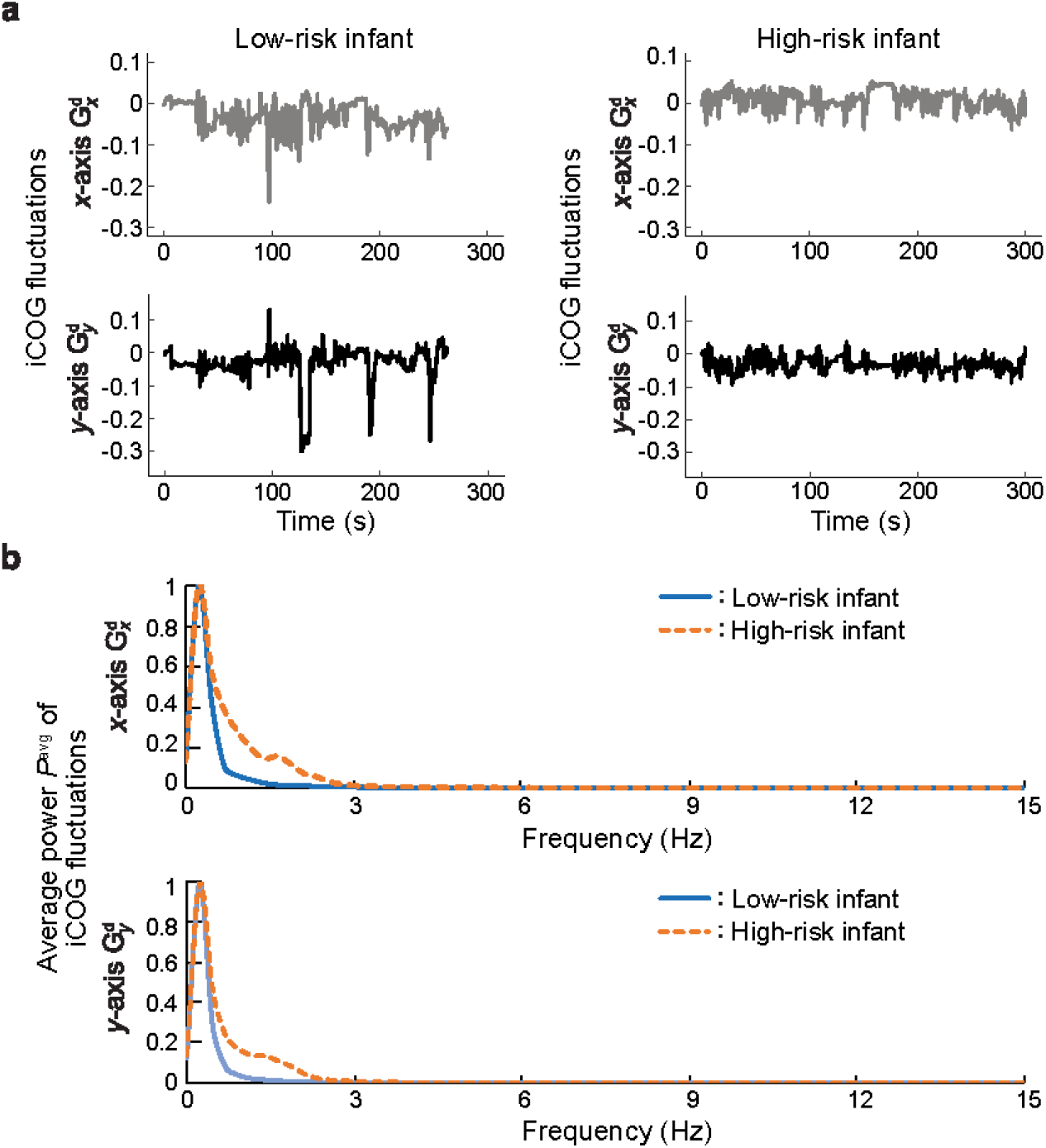
Example data of the temporal course of body center (iCOG) fluctuation (upper panels) and power spectrum of iCOG fluctuation (lower panels).

The standard deviation of the body centre velocity and the fluctuation along the y-axis (*I*_13,*y*_, Stat = −2.03, *p* = 0.0501; *I*_14,*y*_, *W* = −5.59, *p* < 0.001), and the area covered by the trajectory of the body centre excursion (*I*_15_, *W* = −5.96, *p* < 0.001) were significantly smaller in the high-risk group than in the low-risk group.

There were significant (*p* < 0.05) and marginally significant (*p* < 0.10) group differences across the 14 bodily movement features. For each pair out of the 14 features, we computed the correlation coefficient. Correlational coefficients between *I*_9,y_, *I*_12,y_, *I*_11, y_, and between *I*_11, y_ and *I*_11, x_ were above 0.7. Therefore, these four features were averaged into a single feature. The correlation coefficients between *I*_13, x_ and *I*_13, y_, *I*_13, x_ and *I*_15_, *I*_13, y_ and *I*_15_, *I*_14, x_ and *I*_15_, *I*_14, y_, and *I*_15_ were all above 0.7. Thus, these five features were compressed into a single feature as well. After the compression, none of the absolute values of the pairwise correlation coefficients was above 0.7.

### 2-2. Group Classification using Machine Learning Algorithms

Four classifiers were trained using the seven selected features as predictors. The four machine learning algorithms included two linear classifiers, i.e. linear discriminant analysis (LDA) and logistic regression (LR), and two non-linear classifiers, i.e. a multi-layered perceptron (MLP) and a log-linearised Gaussian mixture network (LLGMN; Tsuji et al., 1999). The performance of the MLP and LLGMN depends on the initial state of their random weights. Thus, hyperparameter tuning was repeatedly performed using a nested leave-one-out procedure 10 times, for both the MLP and LLGMN. After the hyperparameter tuning, the average number of nodes in the hidden layer of the MLP was 11.39 (SD = 2.0), while the average number of components in the LLGMN was 2.59 (SD = 1.43). Below, we describe and discuss the performances of the MLP and LLGMN classifiers with the highest F1 score and the highest area under the precision-recall curve (AUC-PR). The confusion matrices generated by the four classifiers are shown in Figure 4. Fisher’s exact test on the data revealed significant associations between classifications made by each of the four classifiers and grouping by the MCHAT score (*ps* < 0.003). Yule’s coefficients of the associations were above 0.85. The performance indicators evaluated using the leave-one-out procedure are summarized in Table 3.

**Figure 4.**
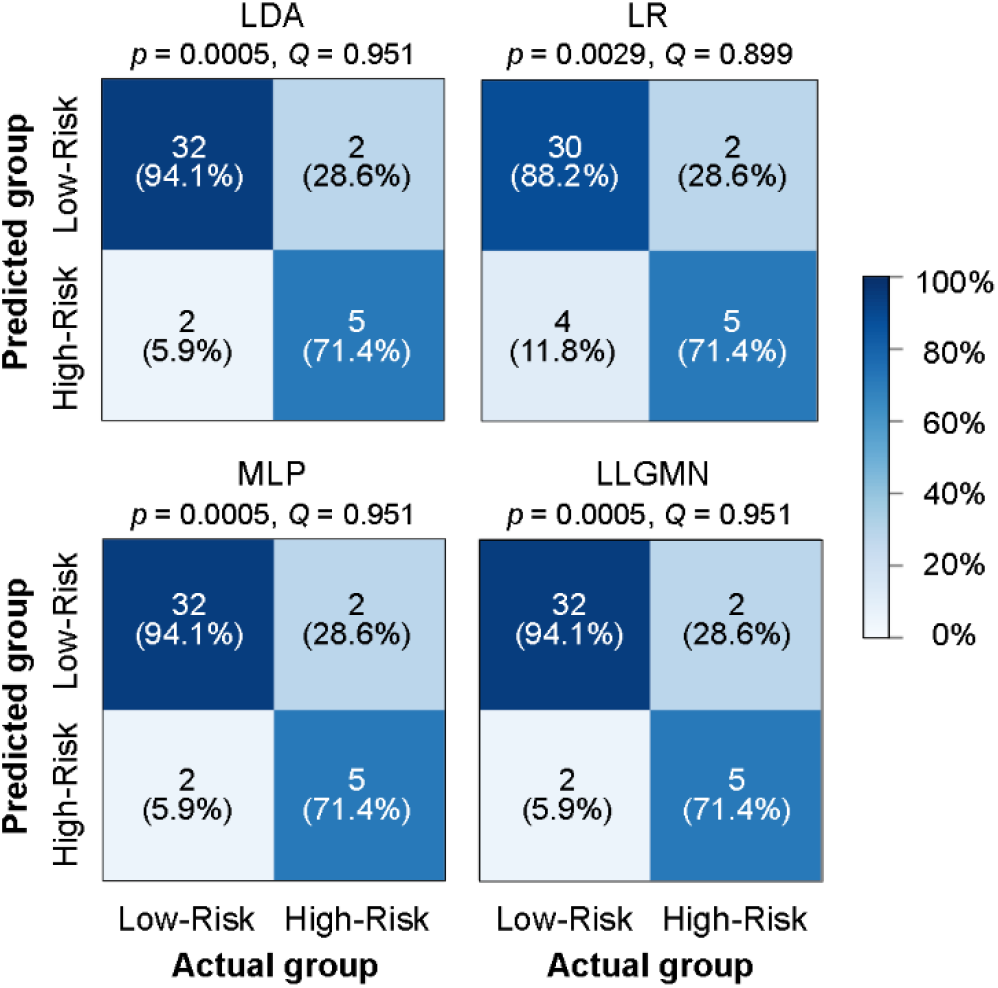
Confusion matrices summarizing the classification results by the four classifiers.

**Table 3.**
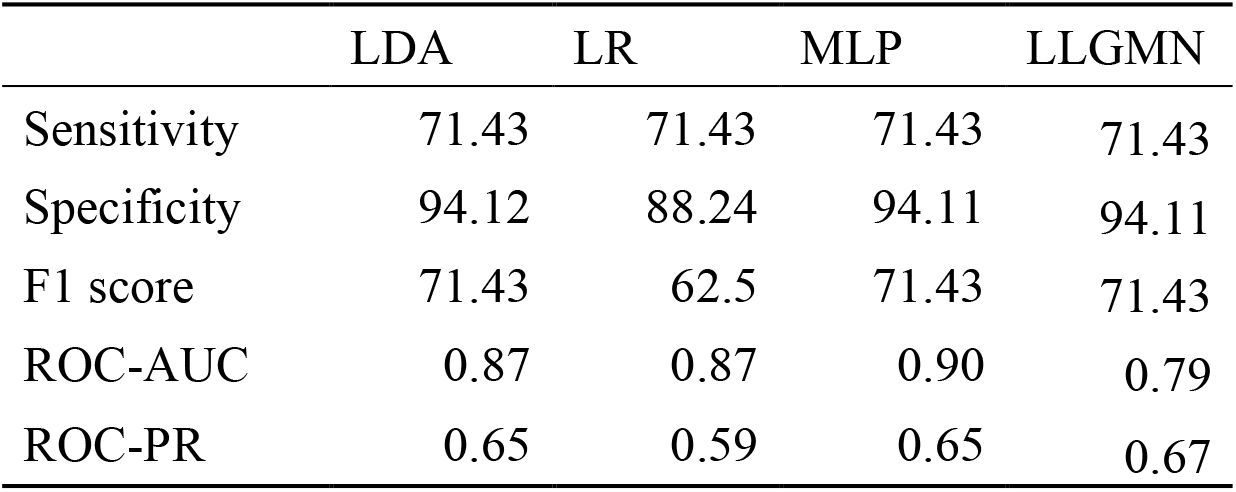
Sensitivity, Specificity, F1 score and AUCs of the four classifiers.

## Discussion

The present study investigated the relationships between automatically extracted features of spontaneous bodily movements of 4-month-old infants and their MCHAT scores at 18 months of age. Infants at risk for ASD showed atypical characteristics for a variety of bodily movement features at 4 months of age. Further, four classifiers with bodily movement features as predictors succeeded in classifying infants with low and high ASD-like behaviours. Taken together, these findings indicate that bodily movement patterns at 4 months of age can inform about ASD-like behavioural tendencies at 18 months of age.

A close look at group differences of bodily movement patterns reveals that the bodily motion patterns of infants with high MCHAT scores are characterised mainly by (i) reduced frequency and strength of lower limb motion, and (ii) lack of rhythmicity in movements. The latter characteristic is reflected in a larger second moment around the central frequency in the time series of the body centre movement for these infants. For low-risk infants, large power was distributed around the central frequency, indicating that these infants moved their body centres periodically within a certain frequency range. Compared with low-risk infants, power was more diffusely distributed for high-risk infants, indicating that their movement patterns lacked notable rhythmic components.

It is generally known that children with ASD exhibit atypical patterns of motor control, such as clumsiness, reduced muscle tone, and poor motor coordination (Paquet et al., 2016; Provost et al., 2007; Fournier et al., 2010). These types of motoric atypicality potentially lead to reduced amplitude and frequency of limb motion and poor coordination of limb movements, as observed in the present study. The lack of rhythmicity in bodily motion patterns might indicate the atypicality of the central pattern generator (CPG) that generates a rhythmic pattern of involuntary movements. Because the CPG is responsible for generating rhythmicity in gait patterns, poor rhythmicity in spontaneous movement patterns during infancy might be linked to atypical gait patterns often observed in children with ASD (Kindregan, Gallagher & Gormley, 2015).

Most items of the MCHAT concern the development of socio-cognitive functions, such as the theory of mind and joint attention. Thus, the present findings nicely dovetail with the observations by Bhat et al. (2012) that motor delay at 6 months of age is associated with delays in social communication at 18 months of age in infants with high risk of ASD. Considering these, one might be tempted to assume that features of spontaneous bodily movement, as quantified in the present study, are linked to later social development. However, it is premature to draw such a conclusion because impairments in motor and socio-cognitive functions might be connected to the atypicality of different sub-regions of identical neural structures in the autistic brain (Qiu et al., 2010).

Several studies have found that early signs of autistic traits can be detected well before the diagnosis of ASD (Adrien et al., 1993; Baranek, 1999; Osterling, Dawson, & Munson, 2002; Klin et al., 2009; Elsabbagh et al., 2012). Consistent with these observations, we succeeded in discriminating infants with high ASD risk from those with low ASD risk, using linear and non-linear classifiers. Overall, the four classifiers exhibited comparable performance, supporting the robustness of our findings that the bodily movement patterns at 4 months of age contain sufficient amount of information for predicting the emergence of ASD-like behaviour at 18 months of age. At the same time, the performance of the two non-linear classifiers was numerically superior to that of the linear classifiers. Astonishingly, the AUC-ROC for the MLP classifier was above 0.9, indicating the possibility that non-linear classification based on bodily movement features can achieve classification performance that is acceptable even in clinical settings.

The present findings provide preliminary results that lead to the establishment of objective markers for the early detection of autistic tendencies in children. A recent surge of studies on digital phenotyping of ASD (Anzulewicz et al., 2016; Dawson et al., 2018; Ardalan et al., 2019) raised the possibility that behavioural features, quantified using low-cost non-invasive sensors, could provide reliable clues for detecting children and adults at risk for ASD. In a closely related study, Anzulewicz et al. (2016) succeeded in classifying children with ASD from their neurotypical counterparts by submitting finger movement patterns captured by touch sensors in tablets to machine learning-based analysis. A similar attempt was also reported by Ardalan et al. (2019) (for a brief review, see Doi, 2020). The current research extends the achievements of these previous studies by raising the possibility that atypicality in bodily movement patterns in infancy could be used for predicting later emergence of ASD-like tendencies during toddlerhood. A series of large-scale clinical studies have shown that ASD can be predicted based on the developmental patterns of brain surface structures (Hazlett et al., 2017) and resting state brain activation (Emerson et al., 2017) measured using magnetic resonance imaging during toddlerhood. It would be of great interest to determine whether comparable performance on the early diagnosis of ASD can be achieved using the digital phenotyping technique adopted in the present study.

The current study had some limitations. First, motor impairment is often observed in other neurological and psychiatric conditions. As such, several researchers questioned the specificity of atypical development of motor function as an early sign of ASD (Ozonoff et al., 2008; Provost et al., 2007). Second, we are not fully confident that bodily movement patterns are linked to ASD, or to the emergence of autism-like behaviours on the sub-clinical level. To address these limitations, the usefulness of our system for early screening of children with ASD should be tested in a long-term study with a prospective design that would recruit children genetically at high risk for ASD.

## 4. Materials and Methods

### 4.1. Participants

Sixty-two 4-month-olds and their mothers participated in video recordings of bodily movement patterns in the present study as a part of the birth cohort study, after the caregivers gave their written informed consents. We used all the data available at the point of analysis without determining the sample size *a priori*, because the number of infants evaluated to be at high risk for ASD was predicted to be quite small. The mother-infant pairs were recruited in Nagasaki City, a rural city on Kyushu Island in Japan, through fliers or in-person advertisement in obstetrics and gynaecology clinics. The protocol of this study was reviewed and approved by the ethics committee of the Graduate School of Biomedical Sciences at Nagasaki University (Registration Number: 14050205-2) and the Graduate School of Engineering at Hiroshima University (Registration Number: E-1150-1).

### 4.2. Procedure

Mother-infant pairs participated in the present study at two time points, i.e. 4 and 18 months after birth. At 4 months of age, the subjects visited our laboratory at Nagasaki University. At this point, we video-recorded the bodily movements of infants while lying in the supine position, for later analysis. Meanwhile, mothers completed several questionnaires regarding their mental state and the development of their infants. We sent out questionnaires, including MCHAT, about a week before each infant reached the age of 18 months after birth, and asked the mothers to send back the questionnaires after answering them fully.

#### 4.2.1. Bodily Movement Recording

Each infant subject was laid on a black mat, in a cream-white wooden crib. The infant’s body, except for the head, arms, and limbs, was covered in a white wrap, for increasing the contrast between the infant’s body and background. A video camera was affixed to a silver stainless bar arching over the crib, and was directed downward toward the infant, as shown in the leftmost panel of Figure 1. The infant movements were recorded continuously. The average length of the recorded videos was 24,028 frames (SD = 8,425 frames; range = 9,600–50,940 frames). The frame rate *f*_s_ and resolution of the video recordings were 30 fps and 720 × 480 pixels, respectively.

#### 4.2.2. Self-Administered Questionnaires

We asked the mothers to complete self-administered questionnaires at three points. The first batch of questionnaires was administered at the time of enrolment. The questionnaires asked for demographic information regarding her and her husband’s age, socio-economic status, and medical history. At 4 months of age, information about the mother’s mental state and infant’s developmental state was collected at the lab; the results of this analysis will be reported elsewhere. At 18 months of age, we mailed questionnaires, including the MCHAT. The MCHAT included 23 items of Yes/No questions, asking whether a child exhibited developmental delays in socio-cognitive functions. The MCHAT also included items about atypical behaviors often observed in ASD, such as hypersensitivity to sensory stimulation. The cut-off point was three.

### 2.3. Analysis

#### 2.3.1. Bodily Movement Analysis

In the initial stage of processing, the recorded videos were divided into video sub-segments by deleting frames that contained the infants’ sleep, crying, and accidental occurrence of external stimulation. Videos of continuous bodily movements were first converted into binary images by image thresholding, to extract the image of the infant’s body as the foreground. Images of spontaneous bodily motion were then obtained by frame-by-frame subtraction. Motion in each area of the body was extracted from these images. The procedure for segmenting a static image into body areas is as follows. First, the outer circumference of the infant’s trunk within each frame was approximated using an ellipsoid. Then, a rectangle was defined such that the rectangle contained an ellipsoid within it with a margin. Thereafter, this rectangle was divided into four sub-regions, each containing the right arm, left arm, right leg, and left leg. Based on the bodily movement data within each quadrant of the entire rectangle, we calculated 26 features of bodily movement, as summarized in Table 2.

#### 4.3.2. Feature Selection

We first selected the features that are useful for classifying infants into low and high ASD risk groups. In the first stage of feature selection, we tested group differences on 26 features between infants with low and high ASD risk, using the Brunner—Munzel test. Features that showed significant group differences at the 10% significance threshold were retained for further analyses. This lenient threshold was adopted because we wished to retain as many informative features as possible for later classification using machine learning algorithms. In the second stage of feature selection, the correlation coefficients were computed for every pair of retained features. Pairs of features with correlation coefficients ≥ 0.7 were lumped into a single feature.

#### 4.3.3 Group Classification by Machine Learning

We examined whether bodily movements contain sufficient information for discriminating infants with high ASD risk from those with low ASD risk, using machine learning approaches. Four classifiers were trained for predicting whether an infant belonged to the high-risk group based on the selected bodily movement features, using supervised learning. The four machine learning algorithms included two linear classifiers, i.e. the linear discriminant analysis (LDA) and the logistic regression (LR), and two non-linear classifiers, i.e. the multi-layered perceptron (MLP) and the log-linearised Gaussian mixture network (LLGMN; Tsuji et al., 1999). The MLP included one hidden layer, the activation function of which was a rectified linear unit. The performance of the four classifiers was cross-validated using the leave-one-out procedure. In the leave-one-out approach, the data of one of the participants were treated as unknown test data, while the data of the remaining participants were used for training the corresponding classifier. This cycle was repeated until the data from every participant served as the test data. The MLP and LLGMN each included one hyperparameter, i.e. the number of hidden layers and the number of components, respectively. These hyperparameters were tuned using a nested leave-one-out procedure (Parvandeh et al., 2020). Fisher’s exact test was performed for testing associations between the classifications made by each of the classifiers used in this study and grouping by the MCHAT scores. Yule’s correlation coefficient (Yule’s Q) was also calculated, for confirming the strength of each association. The performances of the classifiers were compared in terms of the sensitivity, the specificity, the F1 score, the area under curve of receiver-operator-characteristic curve (AUC-ROC), and the area under the precision-recall curve (AUC-PR) as performance indicators.

## Data Availability

All data produced in the present study are available upon reasonable request to the authors

## Acknowledgments

This work was supported by JSPS KAKENHI Grant-in-Aid for Scientific Research(C)(Grant Number 26461769 and 17K01904) to H.D.

## Appendix

### Definitions of Bodily Movement Features

In the bodily movement analysis, each frame of the s-th video sub-segment was converted into a binary image after segregating the infant body from the background. Based on the rectangle defined around the infant body in each frame, seven bodily regions, *A*_*k*_ (*k* = 1–7), were defined. *A*_1∼4_ represented the upper right, upper left, lower right, and lower left quadrants, respectively. *A*_5_ and *A*_6_ were defined as *A*_1_ + *A*_2_ and *A*_3_ + *A*_4_, respectively, thus representing the upper and lower body. Finally, the whole body, *A*_7_, was defined as the sum of *A*_5_ and *A*_6_.

From the binary image, body posture, 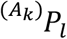, and body movement, 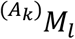, in each area *A*_*k*_ in the *l*-th frame (*l* ≤ *Ls*) were extracted, based on the following equations:

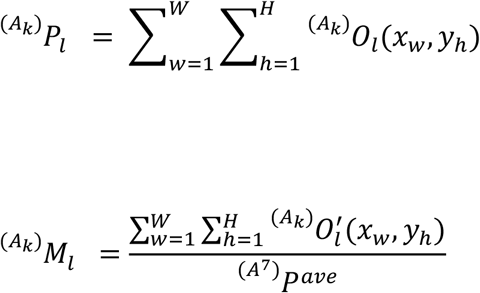

where 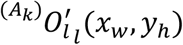 is the pixel value of the image obtained by the frame-by-frame subtraction represented by a binary number (0 or 1) at pixel coordinates (*x*_*w*_, *y*_*h*_) within area 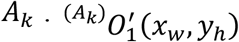 is zero. W and H represent the number of pixels along the x-and y-axes within each area, respectively. 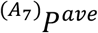 is the average of the maximal values of 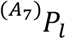 up to the *E*-th frame of the first *Le* (*E* ≤ *Le*) frames of the first video sub-segment. *Le* was set to 30 frames. When 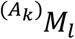 exceeded the threshold value of *M*_*th*_, the *l*-th frame was judged to contain a bodily movement in area *A*_*k*_. We hereinafter refer to a frame that contains a bodily movement in area *A*_*k*_ as “a frame with a bodily movement”

The coordinates of the body centre at the *l*-th frame, (*G*_*l,x*_, *G*_*l,y*_), were calculated by the following equations:

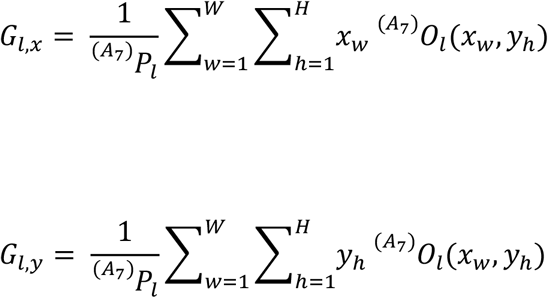

The velocity of the body centre was calculated as the frame-by-frame difference of the body centre coordinates at the *l*-th frame, 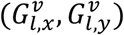. The fluctuation of the body centre 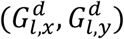 was calculated based on the average of the body centre coordinates, 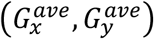, over the first *L*_*e*_ frames of the first video sub-segment, by the following equations:

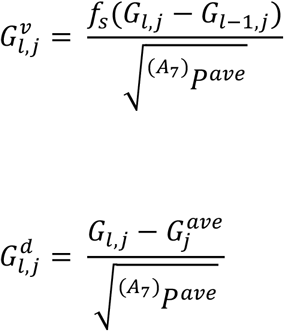

where *j* is *x* or *y*. 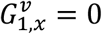 and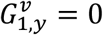.

### 1. Movement Magnitude

#### 1-1. I_1_: Movement Frequency

Movement frequency was defined as the proportion of the number of frames with bodily movement against the total number of frames *L*, which is the total length of video sub-segments:

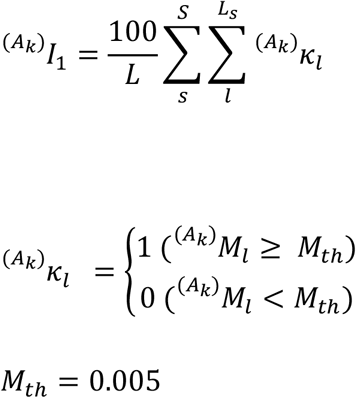

#### 1-2. I_2_: Movement Strength

Movement strength was defined as the average magnitude of movement within frames with a bodily movement. In the following equation, *L*^′^ represents the total number of frames with bodily movements:

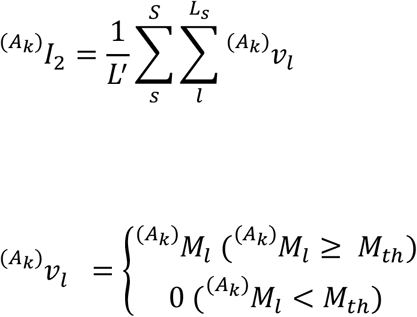

#### 1-3. I_3_: Movement Count

Movement count was defined as the number of temporal segments where continuous bodily movement was detected, divided by the total number of frames *L*. Every frame within each temporal segment met 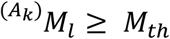. Simultaneously, 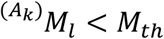 was in the frame just before _*s*_ the first frame of the temporal segment and in the frame immediately after the last frame of the temporal segment. The movement count was calculated by the following equation, where 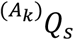 represents the number of temporal segments within the *s*-th video sub-segment:

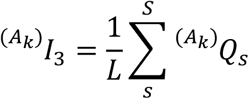

### 2. Movement Balance

#### 2-1. I_4_: Ratio of Movement Frequency

The ratio of the movement frequency between the bodily regions *A*_*k*1_ and *Ak*_*k*2_ was defined by the following equation:

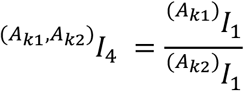

#### 2-2. I_5_: Ratio of Movement Strength

The ratio of the movement strength between the bodily regions *A*_*k*1_ and *A*_*k*2_ was defined by the following equation:

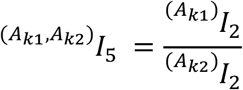

#### 2-3. I_6_: Movement Coordination

Movement coordination between the bodily regions *A*_*k*1_ and *A*_*k*2_ was calculated as the correlation coefficient 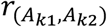 between the time-series data of 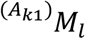 and 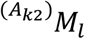 according to the equation below. Correlation coefficients were computed within sliding temporal windows with a length of *L*_*c*_. *L*_*c*_ was set to 300 frames. The stride of the sliding window was one frame. Correlation coefficients were then averaged to yield a single index of the movement coordination.

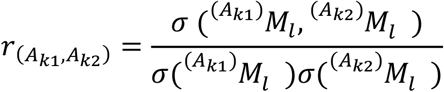

### 3. Movement Rhythm

#### 3-1. Central Frequency (I_7_) and Second Moment around Central Frequency (I_8_) of Movement

The time-series data of *M*_*l*_ of each video sub-segment were analysed in the frequency domain. Within each video sub-segment, the time-series data within a sliding window with the length of *L*_*f*_ were subjected to the fast Fourier transformation. *L*_*f*_ was set to 128 frames. The stride of the moving window was one frame. Then, the average power spectrum density (PSD), *P*(*f*), was computed by grand-averaging PSDs across all the sliding widows in all the video sub-segments. Based on *P*(*f*), central frequency (*F*_*cntr*_) and second moment around the central frequency (*D*_*cntr*_) were computed according to the following equations, where *f*_*max*_ is the maximal frequency range for analysis. *f*_*max*_ was 15 Hz. *F*_*cntr*_ and *D*_*cntr*_, computed on the basis of *P*(*f*), correspond to *I*_7_ and *I*_8_, respectively:

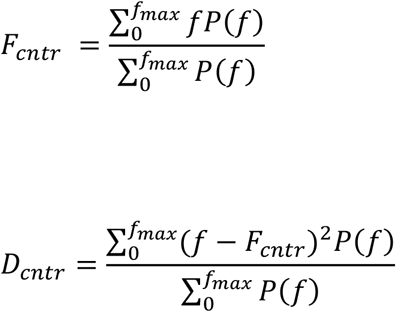

#### 3-2. Central Frequency (I_9,x_, I_9,y_) and Second Moment around Central Frequency (I_10,x_, I_10,y_) of Body Centre Velocity

The time-series data of the body centre velocity along the x- (top-bottom) and y- (left-right) axes were subjected to the fast Fourier transformation. Based on the resulting PSD, the central frequency and the second moment around the central frequency were computed in essentially the same manner as described in ***Section 3-1***.

#### 3-3. Central Frequency (I_11,x_, I_11,y_) and Second Moment around Central Frequency (I_12,x_, M I_12,y_) of Body Centre Fluctuation

The time-series data of body centre fluctuation along the x- (top-bottom) and y- (left-right) axes were subjected to the fast Fourier transformation. Based on the resulting PSD, the central frequency and the second moment around the central frequency were computed in essentially the same manner as described in ***Section 3-1***.

### 4. Movement of the Body Centre

#### 4-1. I_13_: Mean Absolute Body Centre Velocity

The average absolute values of the body centre velocity along the 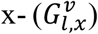 and y-axes 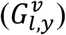 were computed using the following equations:

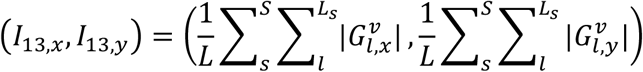

#### 4-2. I_14_: Standard Deviation of Body Centre Fluctuation

The mean standard deviations of the body centre fluctuations along the 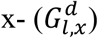 and y- axis 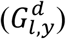, *σ*_*x,y*_, within each video sub-segment were computed by averaging the standard deviations in the sliding temporal windows of length *L*_*g*_, calculated following the equation below. *L*_*g*_was set to 300 frames. The stride of the sliding window was one frame:

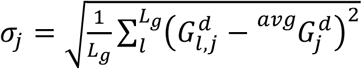

where *j* is *x* or *y*, and 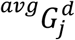 represent the mean of 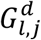 within each sliding window. Then, *I*_14,*x*_ and *I*_14,*y*_ were computed by averaging *σ*_*j*_ along each axis, across all video sub-segments.

#### 4-3. I_15_ : Area of Body Centre Excursion

The area within the outer circumference of the trajectory of the body centre excursion within each video sub-segment was computed by averaging the areas in the sliding temporal windows of length *L*_*g*_. The stride of the sliding windows was one frame. Then, *I*_15_ was computed by averaging the area of body excursions across all video sub-segments.

